# Multi-fidelity Bayesian optimization of population-robust near-infrared sensors for skeletal muscle oximetry

**DOI:** 10.64898/2026.07.08.26357539

**Authors:** Kiran Bhattacharyya

**Affiliations:** SKA Labs, Atlanta, GA, USA

**Keywords:** muscle oximetry, near-infrared spectroscopy, Monte Carlo, Bayesian optimization, multi-fidelity, robust optimization, CVaR, sensor design

## Abstract

Designing transcutaneous skeletal muscle oxygenation (SmO_2_) sensors requires jointly optimizing source–detector geometry and wavelength selection while guaranteeing performance across populations that vary in subcutaneous fat thickness and skin pigmentation. We present a multi-fidelity Bayesian optimization (MFBO) framework that couples Monte Carlo light-transport simulations at two photon-count fidelities to a distributionally robust design objective. An autoregressive Gaussian-process surrogate learns the correlation between inexpensive low-photon-count and accurate high-photon-count simulations, and a cost-aware acquisition function decides both *where* and at *what fidelity* to sample. Robustness across the population is enforced with Conditional Value-at-Risk (CVaR) and entropic-risk (ERM) objectives that target worst-case subjects rather than the population average. On a five-layer forearm tissue model with anthropometric variability we find (i) a fidelity regime that is favorable for MFBO where the low-fidelity surrogate is rank-informative (Spearman *ρ* = 0.84) but biased, at 100× lower cost; (ii) MFBO attains 23% higher robust sensitivity than a strong high-fidelity single-fidelity baseline at equal budget (*p* = 0.035), and avoids the optimistic bias that causes low-fidelity-only optimization to collapse when its designs are validated at high fidelity; (iii) CVaR/ERM objectives improve worst-case tail performance by ~ 23% relative to a mean objective without sacrificing average sensitivity; and (iv) discovered designs improve robust tail sensitivity by roughly 3–6× over commercial and heuristic optode layouts, with the largest gains in the high-fat and high-melanin subpopulations. The methodology bridges stochastic light-transport physics with sample-efficient machine-learning optimization and generalizes to cerebral oximetry, photodynamic therapy planning, and wearable physiological monitors.

## 1. INTRODUCTION

Continuous-wave near-infrared spectroscopy (NIRS) is widely used to monitor skeletal muscle oxygen saturation (SmO_2_) in exercise physiology, sports science, and critical care.^1^ Commercial wearable oximeters such as the Moxy and PortaMon are now in routine research and field use.^2, 3^ A wearable muscle oximeter illuminates the skin with light in the ~ 650–950 nm window and infers the oxygenation of the underlying muscle from diffusely reflected light collected at one or more source–detector separations (SDS). The measurement is fundamentally a depth-discrimination problem: photons must traverse the epidermis, dermis, and a highly variable layer of subcutaneous adipose tissue before probing muscle, and the recovered signal is a pathlength-weighted mixture dominated by whichever layers the chosen geometry and wavelengths happen to interrogate.

Two sources of inter-subject variability dominate performance and confound cross-population deployment. First, subcutaneous adipose tissue thickness (ATT) attenuates and shallows the sampling volume; thicker fat layers systematically reduce the muscle contribution to the signal and blunt the observed dynamic range.^2, 4^ Second, epidermal melanin, which varies by roughly an order of magnitude across Fitzpatrick skin types, adds a strong wavelength-dependent superficial absorber that further biases short-separation channels; the closely related racial bias documented for pulse oximetry underscores how strongly pigmentation can distort optical oximetry.^5^ A sensor geometry that is near-optimal for a lean, lightly pigmented subject can be badly mis-matched for a subject at the opposite extreme, and it is precisely these tail subjects who are most often excluded or mismeasured. Designing for the population *average* is therefore insufficient; the design objective must be explicitly robust to the worst-case subgroups, echoing worst-case and distributionally robust formulations used elsewhere in healthcare and machine learning.^6, 7^

Source–detector separation and wavelength selection are the primary design levers,^8^ but the design space is mixed (continuous separations, discrete wavelength subsets, categorical modality) and the objective, represented here as the oxygenation sensitivity evaluated as an expectation or a tail risk over a population of tissue models, has no closed form. Evaluating a single candidate design requires Monte Carlo (MC) photon-transport simulation across many tissue realizations,^9–12^ and a high-accuracy MC evaluation of one population can take minutes to hours even on a GPU. Exhaustive search or gradient-free heuristics over such an expensive, noisy, mixed-integer objective are impractical.

Bayesian optimization (BO) is the standard tool for sample-efficient global optimization of expensive black-box functions.^13,14^ Multi-fidelity BO (MFBO) extends this by exploiting cheap approximations of the objective (implemented here as MC simulations with far fewer photons) to triage the design space, reserving expensive high-fidelity evaluations for the most promising candidates.^15–18^ MFBO is only advantageous in a specific regime: the low-fidelity model must be informative (well rank-correlated with the truth) yet meaningfully cheaper, and typically biased, so that its predictions must be corrected rather than trusted outright.^17^ Whether transcutaneous NIRS sensor design sits in this regime has not, to our knowledge, been established.

We combine MFBO with distributionally robust objectives to design population-robust SmO_2_ sensors entirely *in silico*. Our contributions are: (1) a characterization of the low-versus high-fidelity MC correlation structure that confirms this problem lies in the regime where MFBO is appropriate; (2) a controlled comparison showing MFBO outperforms strong single-fidelity BO baselines at equal computational budget, and, critically, that low-fidelity-only optimization is deceptively optimistic; (3) evidence that tail-robust Conditional Value-at-Risk (CVaR) and entropic-risk (ERM) objectives materially reshape designs to improve worst-case performance; and (4) a demonstration that the discovered designs substantially outperform commercial and heuristic optode layouts, especially for the hardest subpopulations. We frame the case for MFBO as the consensus view of that literature; empirical claims are backed by the experiments in Section 3.

## 2. METHODS

### 2.1 Five-layer Tissue Model and Population Variability

We model the forearm/calf as a planar five-layer turbid medium (Fig. 1): epidermis (0.08 mm), dermis (1.5 mm), subcutaneous fat (2–15 mm), fascia (0.75 mm), and a semi-infinite skeletal muscle layer that is the SmO_2_ target. Each layer is assigned wavelength-dependent absorption from its chromophore composition (oxy- and deoxy-hemoglobin, myoglobin, water, lipid, and, in the epidermis, melanin) and reduced scattering following a power law 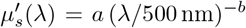.^8^ Melanin absorption in the epidermis uses the standard power-law model *µ*_*a*_ ∝ (*λ/*694)^−3.33^.^8^

**Figure 1.**
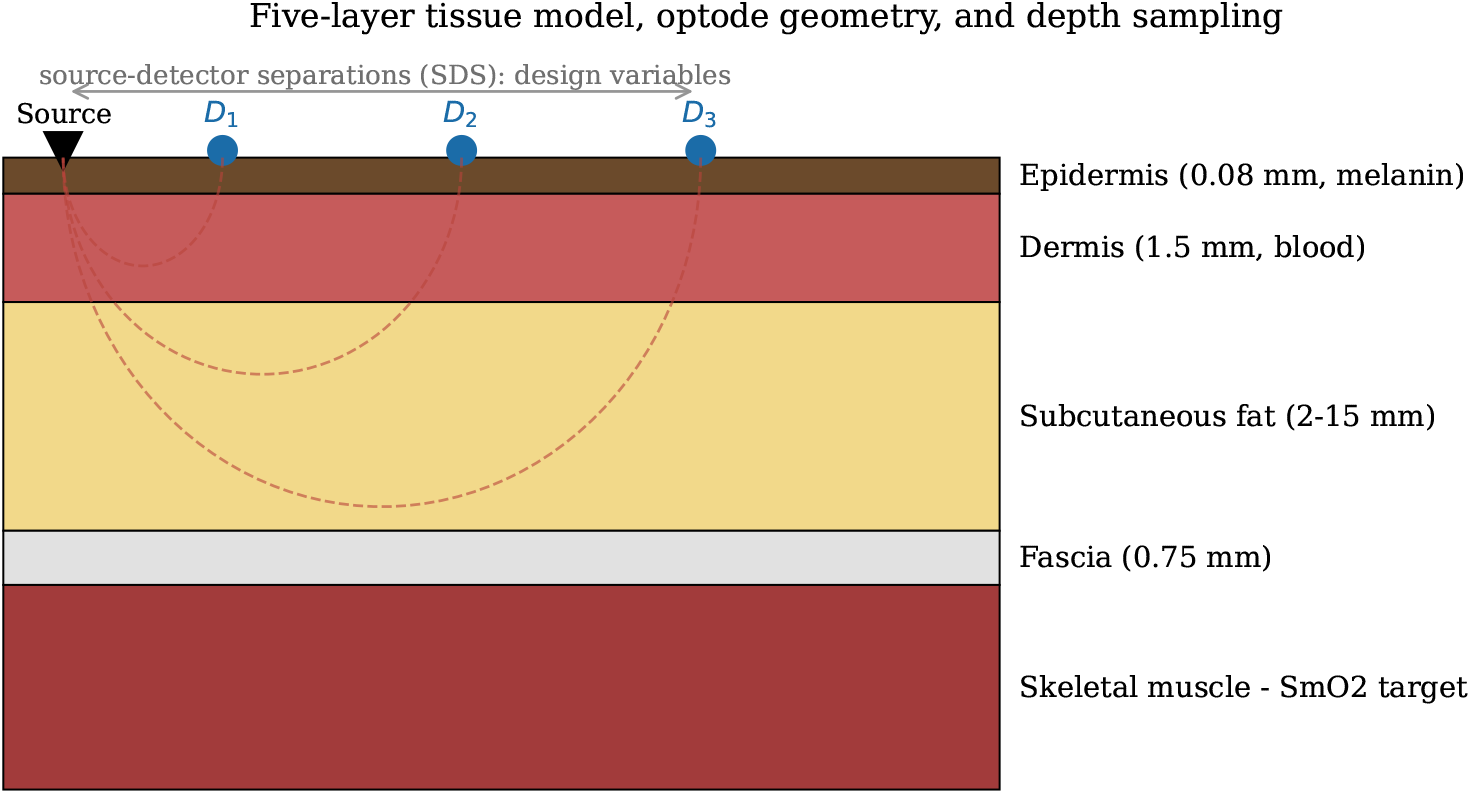
Five-layer tissue model, linear multi-distance optode geometry, and banana-shaped depth-sampling regions for short, medium, and long source–detector separations. Epidermal melanin and the variable subcutaneous fat layer are the dominant sources of inter-subject variability; the three separations *D*_1_, *D*_2_, *D*_3_ and the selected wavelengths are the design variables.

Inter-subject variability is represented by sampling a population of tissue realizations from physiologically motivated distributions: fat thickness log-normal (mean 5 mm, clipped to [2, 15] mm), epidermal melanin volume fraction uniform on [0.01, 0.10] (spanning Fitzpatrick I–VI), and normal distributions for dermal/muscle blood volume, dermis thickness, and muscle myoglobin concentration. Baseline muscle oxygenation is SmO_2_ = 70%. Each population draw defines one “subject” at which any candidate design can be evaluated; robustness objectives (Section 2.3) are computed over these subjects.

### 2.2 Monte Carlo Light Transport and Fidelity Levels

Photon transport through each tissue realization is simulated with a GPU-accelerated voxel-based Monte Carlo engine (MCX,^11^ accessed through its Python binding pmcx). A pencil-beam source illuminates the surface and detected weight is accumulated at ring detectors placed at the design separations. For each design and subject we run a baseline simulation at SmO_2_ = 70% and a perturbed simulation at SmO_2_ = 75%; layer-resolved partial pathlengths and detected reflectance are recorded for the sensitivity calculation (Section 2.3). Continuous-wave (amplitude) and frequency-domain (phase/AC) observables are both supported; the experiments reported here use the continuous-wave modality that dominates commercial wearables.

*Fidelity* is controlled by the launched photon count. The low-fidelity (LF) level uses 5 × 10^4^ photons and the high-fidelity (HF) level 5 × 10^6^ photons, a 100 × ratio in cost that trades stochastic variance for speed. We assign relative computational costs *c*_LF_ = 1 and *c*_HF_ = 100 used by the acquisition function (Section 2.5).^*^

### 2.3 Design Space and Oxygenation Sensitivity

A design *x* comprises three continuous source–detector separations (short ∈ [8, 18], medium ∈ [20, 32], long ∈ [34, 45] mm), a selection of four wavelengths from the candidate set { 660, 690, 730, 760, 800, 830, 870, 910, 940} nm (encoded as binary indicators), a categorical modulation frequency, and a categorical measurement modality. The primary per-subject figure of merit is the continuous-wave oxygenation sensitivity, the magnitude of the change in log-reflectance per unit change in muscle oxygenation,

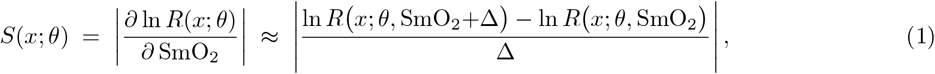

where *θ* denotes the sampled tissue parameters of a subject, *R* is the detected diffuse reflectance, and Δ = 0.05. When multiple wavelengths and detectors are present the per-channel sensitivities are combined by a root-mean-square aggregation across wavelengths and the best-performing detector is used; higher *S* indicates a design better able to resolve muscle oxygenation changes against the tissue background.

### 2.4 Distributionally Robust Objectives

Let *S*(*x*; *θ*) be the sensitivity for design *x* at subject *θ*, and let the population induce a distribution over *S*. We compare three population objectives, all maximized:

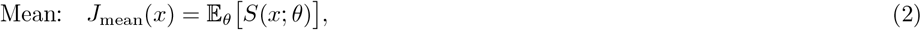

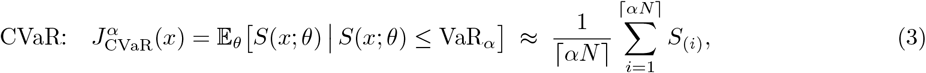

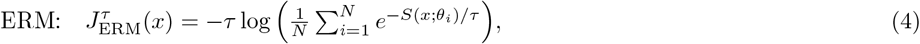

where *S*_(1)_ *S*_(*N*)_ are the order statistics over *N* population samples and *α* = 0.1 selects the worst 10%. CVaR is the standard coherent tail-risk measure:^19^ maximizing it lifts the average of the worst-off subjects. CVaR and related risk measures have recently been adopted as objectives within Bayesian optimization of expensive black-box functions.^20^ The entropic-risk measure (ERM) is a smooth alternative that we introduce as a drop-in objective; it is a coherent risk measure that uses *all N* samples (rather than only the worst #x2308; *αN* ⌉), giving an *N*-fold less noisy gradient signal for the surrogate while retaining tail emphasis. ERM nests the mean (*τ* → ∞) and the worst case (*τ* → 0); we use *τ* ≈ 1.5–2, comparable to CVaR at *α* ≲ 0.05. CVaR/ERM also expose the worst-case subject indices, which the optimizer can target for focused high-fidelity evaluation.

### 2.5 Multi-fidelity Bayesian Optimization

We place an autoregressive (AR1) multi-fidelity Gaussian-process (GP) prior on the objective.^15^ The high-fidelity objective is modeled as a scaled low-fidelity process plus an independent discrepancy,

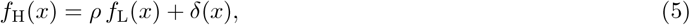

where *f*_L_ and *δ* are independent GPs with Matérn/RBF-type kernels and *ρ* is a learned cross-fidelity correlation coefficient (estimated from paired observations and clipped to [0.1, 0.99]). This structure lets abundant cheap LF data shape the surrogate while sparse HF data correct its bias. Candidate designs are proposed by maximizing a cost-aware multi-fidelity Expected Improvement acquisition,

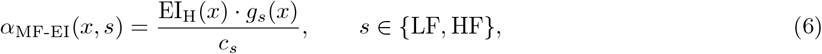

where EI_H_ is the expected improvement in the high-fidelity objective, *c*_*s*_ is the fidelity cost, and *g*_*s*_ is an information-gain factor (unity at HF; 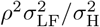 at LF) that credits a low-fidelity query with the fraction of high-fidelity uncertainty it is expected to remove. At each iteration the design–fidelity pair maximizing Eq. (6) is evaluated, the GP is updated, and its hyperparameters (including *ρ*) are re-estimated, until the computational budget is exhausted. Optimization begins from a Latin-hypercube design of experiments with LF and HF initial points.

Single-fidelity baselines are obtained by forcing the same pipeline to a single level: **SFBO-HF** evaluates only high-fidelity simulations (accurate but few evaluations per budget), and **SFBO-LF** evaluates only low-fidelity simulations (many cheap but biased evaluations).

### 2.6 Experimental Protocol

All optimization runs used a per-run budget of 500 LF-equivalent cost units, a population of 20–30 tissue samples for objective estimation during search, *α* = 0.1 for CVaR, and were repeated over 15–40 random seeds depending on the experiment to quantify run-to-run variability. Reported “final” performance and cross-objective comparisons are computed by re-evaluating the best design of each run on a held-out test population at high fidelity (100–500 subjects), which removes optimization-time noise and, for SFBO-LF, exposes the true (unbiased) performance of its LF-selected designs. Statistical comparisons use paired *t*-tests and Wilcoxon signed-rank tests across seeds with Cohen’s *d* effect sizes. Table 1 summarizes the configuration.

**Table 1.**
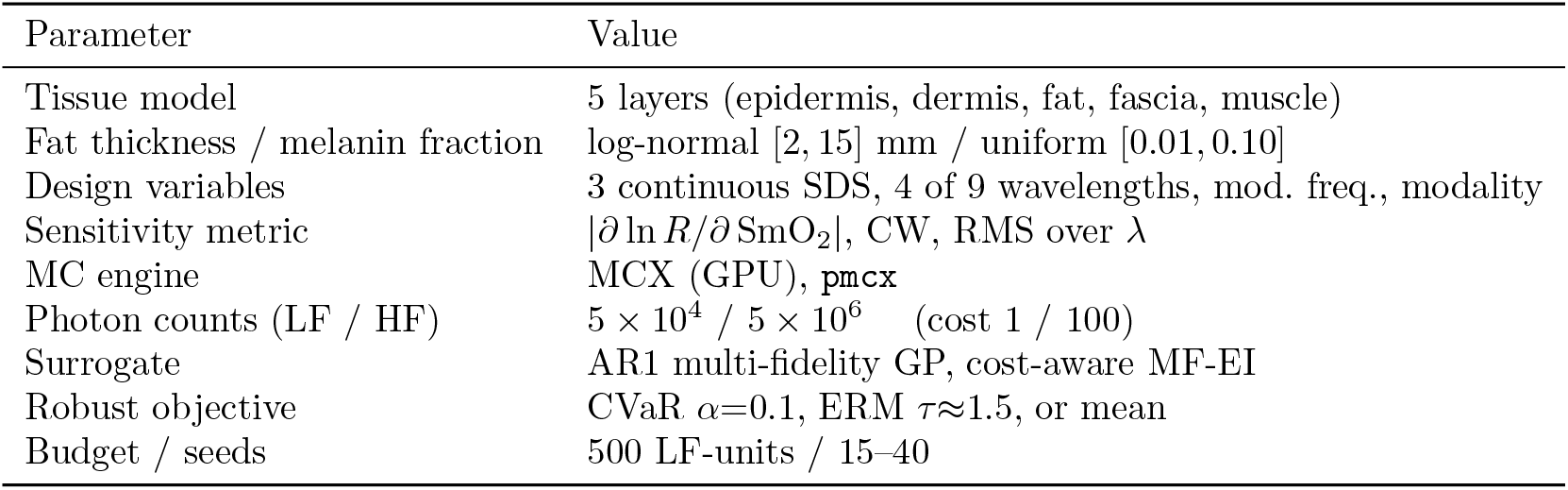
Key configuration for the reported experiments.

## 3. RESULTS

### 3.1 The Problem Sits in a Regime Favorable for Multi-fidelity Optimization

MFBO only helps when the cheap model is rank-informative but biased. We evaluated 150 designs at both fidelities on a common population and compared the paired CVaR values (Fig. 2). The low- and high-fidelity objectives are strongly *rank*-correlated (Spearman *ρ* = 0.84, Kendall *τ* = 0.64, both *p <* 10^−20^) yet only moderately correlated in value (Pearson *r* = 0.60, *R*^2^ = 0.35), with a fitted slope of 0.50 and a non-negligible, heteroscedastic bias (mean − 0.35, s.d. 2.3, up to 8.3 sensitivity units for individual designs). In other words, low-photon-count simulations reliably tell which designs are *better* but not by *how much*, and they cannot be trusted as an unbiased estimate of the truth. Combined with the 100 × cost ratio, this is precisely the informative-but-biased, cost-separated regime in which multi-fidelity surrogates are expected to pay off,^17^ and it motivates the AR1 model’s explicit *ρ*-scaling and additive discrepancy.

**Figure 2.**
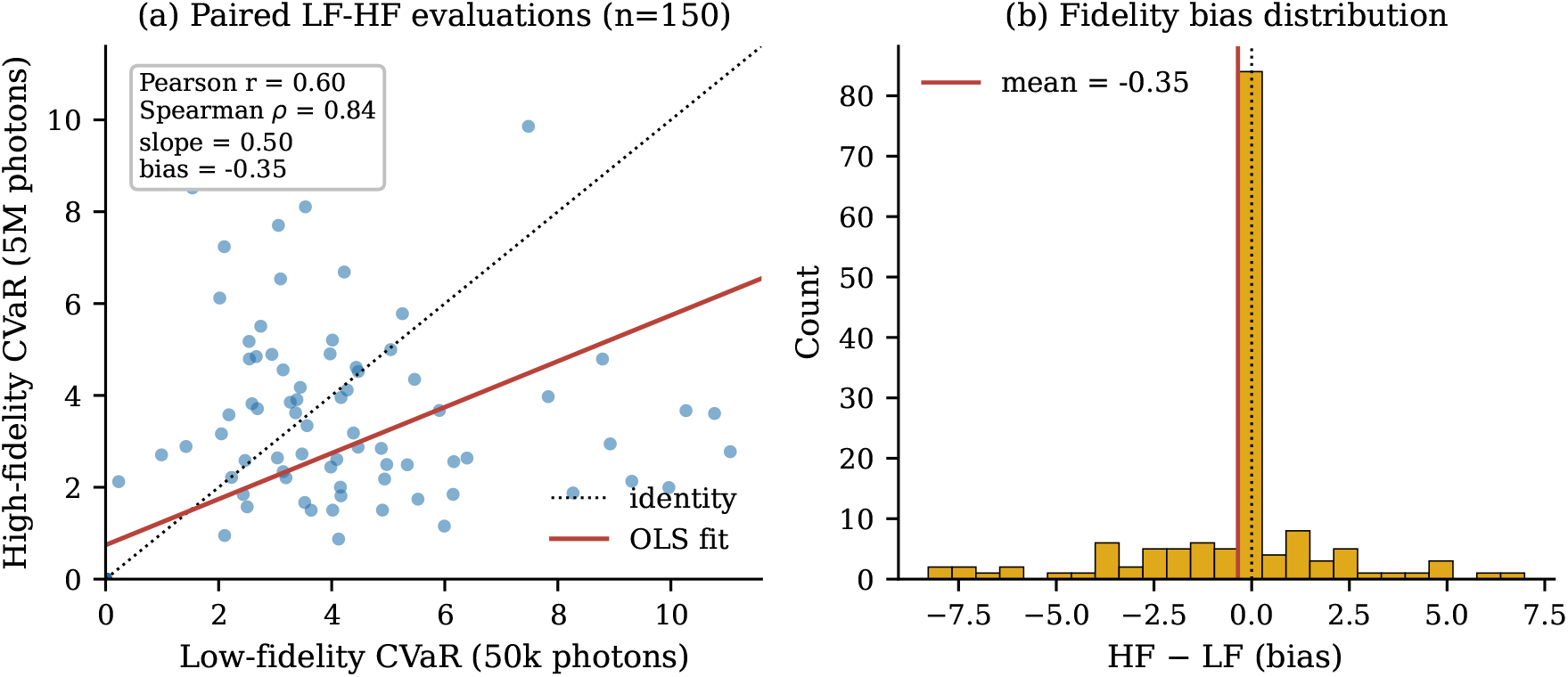
Low-versus high-fidelity agreement over 150 paired design evaluations. (a) Paired CVaR values: strong rank correlation (Spearman *ρ* = 0.84) but only moderate value correlation (Pearson *r* = 0.60) and a fitted slope of 0.50 relative to the identity line (dotted). (b) The fidelity bias (HF*−*LF) is centered near zero but broad, confirming that the low-fidelity model is informative yet biased. This is preciesely the regime in which multi-fidelity optimization is advantageous.

### 3.2 MFBO Outperforms Single-fidelity Baselines at Equal Budget

Across 15 seeds at a fixed budget of 500 cost units, MFBO reached a mean final CVaR of 8.23 ± 1.90, versus 6.70 ± 1.29 for the high-fidelity-only baseline SFBO-HF resulting in a 22.8% improvement that is statistically significant (paired *t*-test *p* = 0.035; Wilcoxon *p* = 0.026; Cohen’s *d* = 0.94, a large effect). The convergence traces (Fig. 3a) show why: at equal budget SFBO-HF can afford only a handful of expensive evaluations and remains starved of data, whereas MFBO uses cheap low-fidelity queries to explore broadly and spends its scarce high-fidelity budget on refinement, dominating SFBO-HF at every budget level.

**Figure 3.**
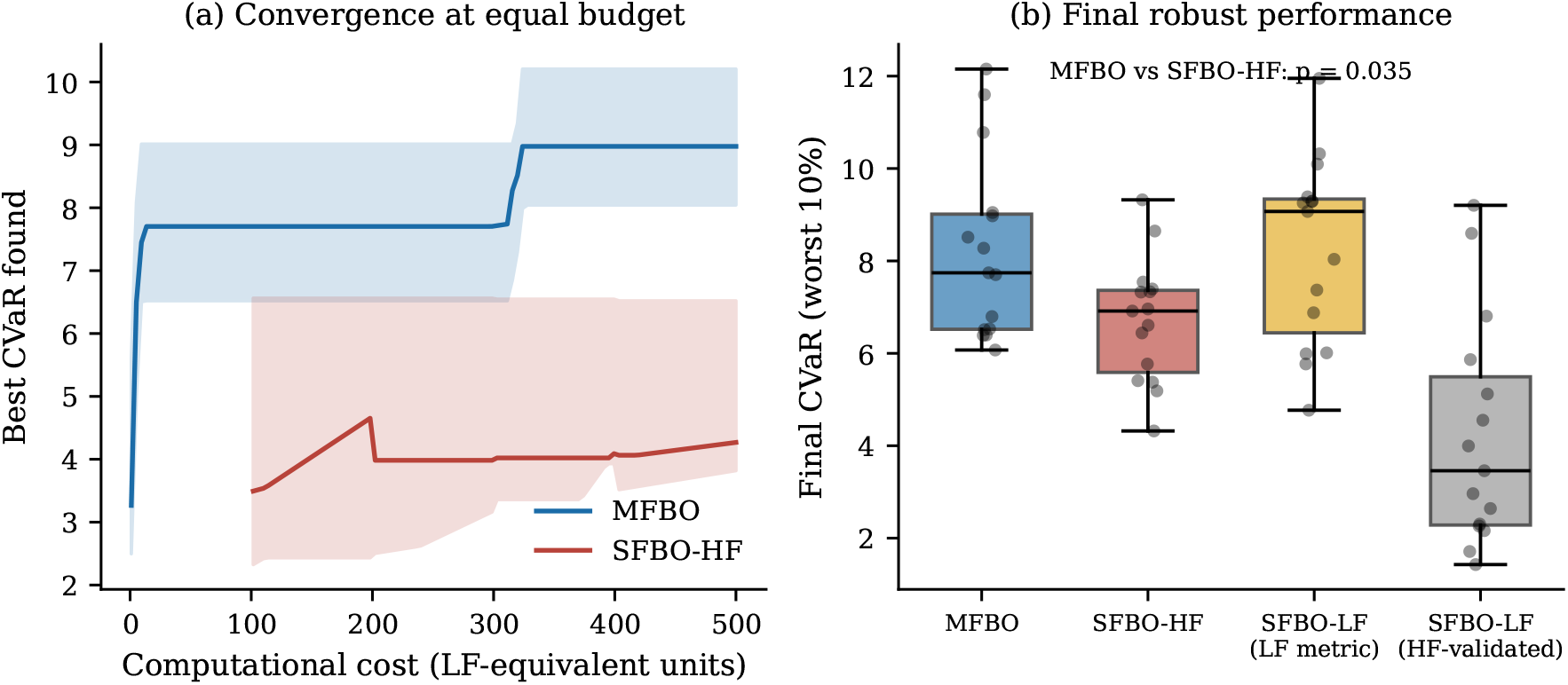
MFBO versus single-fidelity baselines (15 seeds, equal 500-unit budget). (a) Median convergence (bands: interquartile range) of the best CVaR found versus computational cost; MFBO dominates high-fidelity-only BO throughout. (b) Final robust performance. MFBO significantly exceeds SFBO-HF (*p* = 0.035). SFBO-LF looks competitive on its own optimistic low-fidelity metric (third box) but collapses when its designs are validated at high fidelity (fourth box), illustrating the danger of trusting cheap simulations alone.

The comparison with the low-fidelity-only baseline SFBO-LF is more subtle and, we argue, the more important cautionary result. Judged on its own low-fidelity simulations of 50k photons, SFBO-LF appears to match MFBO judged on the high-fidelity simulations on 5M photons (8.23 vs 8.23; Fig. 3b, third box). But its optimization metric is biased and optimistic: when the designs SFBO-LF selected are re-evaluated at high fidelity, their true CVaR collapses to 4.20 ± 2.38 (fourth box) which is 49% below its own reported value and 49% below MFBO (paired *t*-test *p* = 2.5 × 10^−4^, Cohen’s *d* = 1.81). Low-fidelity-only optimization thus overfits to photon-noise and low-count artifacts, “discovering” designs that do not survive honest evaluation. MFBO avoids this failure mode because its AR1 surrogate anchors the cheap data to a smaller number of high-fidelity observations (Table 2).

**Table 2.** Final robust performance (CVaR, worst 10%) across 15 seeds. Higher is better. SFBO-LF is shown both on its own low-fidelity metric and after high-fidelity validation. MFBO and SFBO-HF metrics shown are from high-fidelity validation only.

| Method | Mean ± s.d. | vs. MFBO | Significance |
| --- | --- | --- | --- |
| MFBO (ours) | $8.23 \pm 1.90$ | — | — |
| SFBO-HF | $6.70 \pm 1.29$ | −18.6% | $p=0.035$, $d=0.94$ |
| SFBO-LF (LF metric) | $8.23 \pm 1.96$ | $\approx 0\%$ | n.s. (biased metric) |
| SFBO-LF (HF-validated) | $4.20 \pm 2.38$ | −48.9% | $p=2.5 \times 10^{-4}$, $d=1.81$ |

### 3.3 Robust Objectives Reshape Designs and Improve the Tail

We next asked whether optimizing a robust objective, rather than the population mean, changes the resulting designs and their worst-case performance. Across 40 seeds we ran the optimizer with the mean, CVaR, and ERM objectives and evaluated every resulting design on a common high-fidelity test population, scoring each on mean sensitivity and on the tail metrics (Fig. 4). Relative to the mean objective, the CVaR objective improved worst-10% CVaR by 23.0% and the ERM objective by 22.9% (paired Wilcoxon *p* ≈ 0.03–0.05); worst-5% CVaR and the outright minimum improved comparably. Notably, the robust objectives achieved this *without* sacrificing average performance since mean sensitivity was also 18–20% higher. This indicates that on this problem the mean objective tends to overfit the small optimization-time population and generalizes worse than the robust objectives, which implicitly regularize toward broadly-performing designs. The per-seed improvement distribution is positively skewed (Fig. 4): on most seeds the objectives yield similar designs, but on a substantial minority the mean objective selects a fragile design that the robust objectives repair, producing large tail gains. ERM matched CVaR while using the full population each iteration, consistent with its lower-variance design rationale.

**Figure 4.**
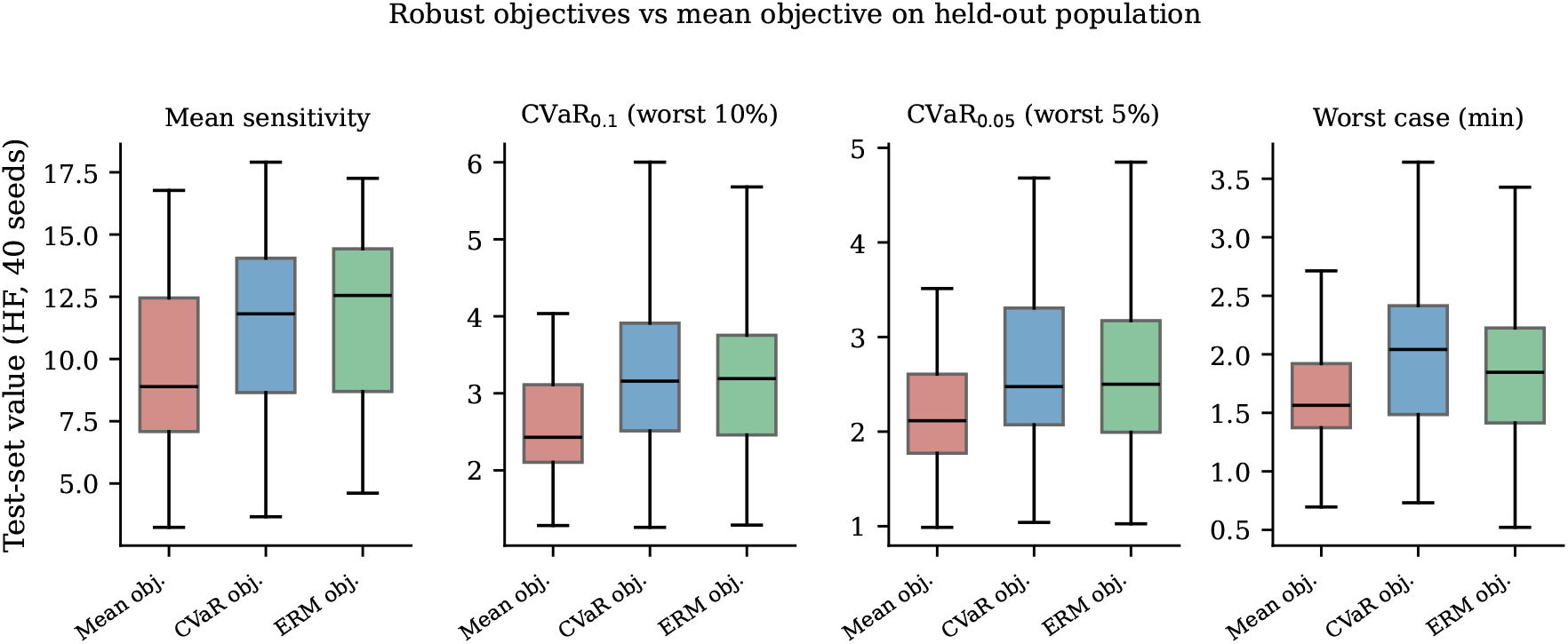
Held-out high-fidelity performance of designs found under the mean, CVaR, and entropic-risk (ERM) objectives (40 seeds). CVaR and ERM improve the worst-case tail metrics (CVaR_0.1_, CVaR_0.05_, min) by *~* 23% relative to the mean objective while also modestly improving mean sensitivity, showing that the robust objectives lift the tail without a mean-performance penalty on this problem.

### 3.4 Discovered Designs Beat Commercial and Heuristic Layouts

Finally we compared a CVaR-optimized design against three fixed heuristic optode layouts representative of practice: a PortaMon-like spatially-resolved arrangement (30*/*35*/*40 mm, 760*/*850 nm), a Moxy-like short-separation wearable (12.5*/*25 mm, four wavelengths), and a generic short-separation layout (10*/*20*/*30 mm, 760*/*850 nm).^2, 3^ All designs were evaluated on a common 500-subject high-fidelity test population (Fig. 5). The discovered design had separations ≈ 13*/*31*/*34 mm with wavelengths 660*/*690*/*870*/*940 nm and improved population mean sensitivity by 20%, 49%, and 151% over the PortaMon-like, short-separation, and Moxy-like layouts respectively, but the gains in *robust* sensitivity were far larger: worst-10% CVaR improved by roughly 2.9 ×, 3.5 ×, and 5.6 × (i.e. 194–457%). The advantage was concentrated exactly where it matters, in the hardest subpopulations: within the highest-fat and highest-melanin quartiles the heuristic layouts nearly lose the muscle signal (CVaR approaching zero), whereas the optimized design retains substantial tail sensitivity (Fig. 5b).

**Figure 5.**
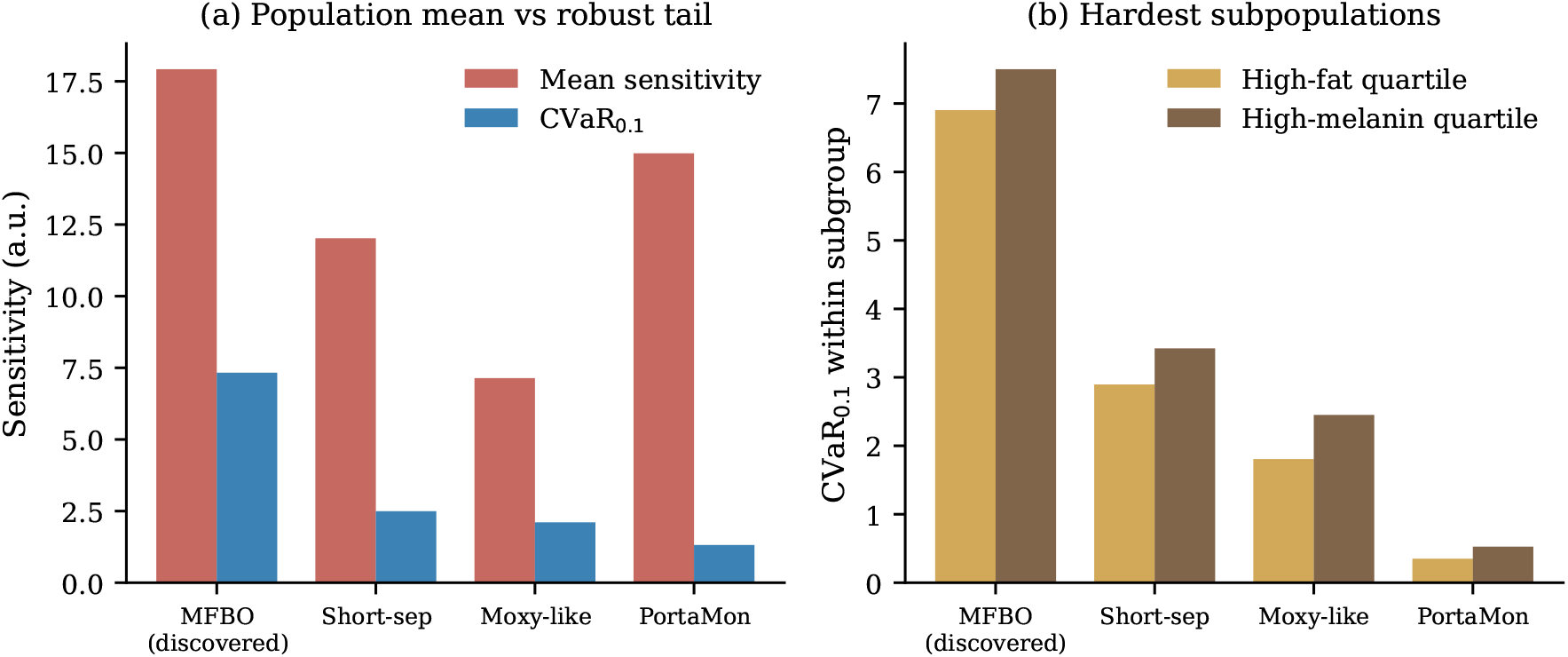
Discovered (CVaR-optimized) design versus commercial/heuristic optode layouts on a common 500-subject high-fidelity test population. (a) The discovered design improves both mean and worst-10% (CVaR) sensitivity, with the robust-tail gain (*~* 3–6*×*) far exceeding the mean gain. (b) Within the hardest subpopulations (high-fat and high-melanin quartiles) the heuristic layouts nearly lose the muscle signal, while the optimized design preserves tail sensitivity.

## 4. DISCUSSION

Taken together, the four experiments form a coherent argument for how to design population-robust NIRS sensors efficiently. Section 3.1 establishes that low-photon-count MC simulations are rank-informative but biased at 100 × lower computational cost. These are the necessary preconditions for multi-fidelity methods. Section 3.2 shows MFBO converts that structure into a concrete advantage, beating a strong high-fidelity BO baseline by 23% at equal budget, and exposes a failure mode that matters for anyone tempted to optimize purely on cheap simulations: low-fidelity-only optimization is optimistically biased and its designs degrade sharply under honest evaluation. This is a practical warning for *in silico* sensor design, where the temptation to use fast approximate forward models is strong. Section 3.3 shows that the choice of *objective*, not just the optimizer, shapes the outcome: tail-focused CVaR/ERM objectives improve worst-case subjects and, on this problem, do so essentially for free in mean performance. This is the right target when the goal is equitable performance across skin tones and body compositions. Section 3.4 closes the loop by showing the discovered designs dominate real-world heuristic layouts by several fold in the tail, precisely for the subjects current devices serve worst.

Several limitations temper these conclusions and should be addressed before translation. The forward model is a planar, five-layer approximation with literature-derived optical properties and assumed population distributions; absolute sensitivity values and the exact optimal geometry will shift with more realistic anatomy (curvature, heterogeneity, vasculature) and with population priors calibrated to a target cohort.^8^ The “sensitivity” objective in Eq. (1) is a forward-model contrast metric and not a full estimator of oxygenation accuracy; incorporating a realistic noise model and an inversion step would tighten the link to end-to-end measurement error. The AR1 surrogate assumes a single global cross-fidelity correlation, and a nonlinear or input-dependent *ρ* (e.g. NARGP^21^) could better capture the heteroscedastic bias seen in Fig. 2. Finally, the cross-objective differences in Section 3.3, while consistent, are modest per-seed and driven by a minority of runs; larger seed counts and matched optimization/test populations would sharpen the estimate. We also note that our positive framing of MFBO reflects the consensus of the multi-fidelity literature; a skeptic could argue that the equal-budget advantage over SFBO-HF depends on the chosen budget and cost ratio, and that at very large budgets the methods should converge. This is a regime we did not probe.

## 5. CONCLUSION

We presented an in-silico framework that couples multi-fidelity Bayesian optimization with distributionally robust objectives to design population-robust skeletal-muscle oximetry sensors. On a five-layer tissue model with anthropometric variability, the framework operates in a fidelity regime favorable to MFBO, outperforms strong single-fidelity baselines at equal budget while avoiding the optimistic bias of cheap-only optimization, produces designs whose worst-case performance is materially improved by CVaR/ERM objectives, and yields optode layouts that beat commercial heuristics by roughly 3–6 × in robust tail sensitivity. The largest gains were for high-fat and high-melanin subjects. The approach is directly transferable to cerebral oximetry,^22^ photodynamic-therapy planning, and other wearable physiological monitors where expensive stochastic forward models and population heterogeneity must be handled together.

## Data Availability

All data produced in the present study are available upon reasonable request to the authors

## Footnotes

* The software supports an intermediate fidelity and larger photon budgets; the two-fidelity setting above was used for all optimization experiments in this paper to keep the fidelity structure interpretable.

## Notes

### Competing Interest Statement

The authors have declared no competing interest.

